# Serum IL-6, IL-10, TNF-α and IFN-γ bio-signature for neonatal sepsis diagnosis and treatment outcome

**DOI:** 10.1101/2025.02.12.25320841

**Authors:** Suraj Thottathil, Priyakshi Gogoi, Vrinda Satheesan Nair, Ambalika Roy, Riya Ahmed, Poonam Dagar, Pradeep Debata, Rajni Gaind, Gita Prashad Kaushal, Renu Gur, Kirti Nirmal, Ravinder Kaur, Sushma Nangia, Vivek Kumar, Anmol Chandele, Krishnamohan Atmakuri, Kajal Jain, Ramesh Agarwal, Mari Jeeva Sankar, Dipankar Ghosh, Ranjan Kumar Nanda

## Abstract

Diagnosing sepsis in preterm neonates is a significant challenge, underscoring the urgent need for timely and accurate methods. In this study, preterm neonates (n=252, 25–35 weeks’ gestation, female: 40%) from sepsis–suspected (n=188) and healthy controls (n=64) were enrolled. Based on the microbiology and mass spectrometry data, the sepsis suspects were categorized into culture-positive/negative (CP/CN) or no sepsis (NS). Serum inflammatory proteins were profiled using the Olink® Target 96 inflammation panel and validated in independent sets using ELISA, multiplex cytokine arrays and qRT-PCR. Serum trace elements (⁵⁷Fe, ⁶⁶Zn, ⁶³Cu, ⁷⁷Se, ⁴⁴Ca, ^24^Mg) were profiled using inductively coupled plasma mass spectrometry (ICP-MS). An increased IL-6: IFN-γ ratio was observed in CP and CN, which normalised upon recovery. Elevated serum IL-6, TNF-α, and IL-10 levels were observed in the CP sepsis groups with respect to CN. A reduced serum IFN-γ level was observed in the CP, CN and NS groups. The qRT-PCR of *IL6*, *TNFA,* and *IL10* showed higher expression in the Cd45+ leukocytes of CP and CN compared to the HC groups, which decreased upon recovery. Serum iron levels were significantly lower in the CP group, indicating anaemia of inflammation, which normalised upon completion of the therapeutic regimen. Serum inflammatory proteins showed group-specific trends, and the serum biosignatures of IL-6, IL-10, IFN-γ, and TNF-α could be useful for distinguishing sepsis subtypes and for diagnosing and monitoring therapeutic responses in neonatal sepsis.

## Introduction

Neonatal sepsis is a severe and complex systemic immune response to infections caused by bacteria, viruses, or fungi, leading to significant morbidity and mortality.^1^ The World Health Organisation (WHO) reported 2.3 million neonatal deaths annually, with sepsis being the key contributor.^2^ The clinical presentation is often non-specific, with symptoms including temperature instability, respiratory distress, feed intolerance, lethargy, and, in severe cases, multi-organ dysfunction.^3^ Accurate and timely classification of sepsis suspects to culture- positive (CP), culture-negative (CN), and no-sepsis (NS) cases remains a significant challenge. Current diagnostic methods primarily rely on laboratory-based microbiology culture techniques for pathogen identification, supplemented by advanced tools such as MALDI-TOF mass spectrometry and multiplex PCR. Other diagnostic approaches include assessing multiple markers, such as inflammatory proteins, micronutrient levels, white blood cell counts, and cytokines. Blood culture tests yield results in 24-48 hours and may yield false negatives in cases of prior antibiotic exposure. However, accurate and early diagnosis requires multidimensional strategies that integrate clinical symptoms with biomarkers to enable timely treatment and avoid unnecessary antibiotic use.^4,5^ Within hours of infection, the inflammatory response begins, and identifying the inflammatory markers may provide a more comprehensive approach to sepsis diagnosis.^6,7^ Previous studies have highlighted that identification of a combination of biomarkers rather than individual ones like CRP or IL-6 would increase the specificity in differentiating neonatal sepsis cases from the healthy control.^8,9,10^ This highlights the urgent need for reliable biosignatures those can accurately distinguish the sepsis subtypes and thereby improve clinical decision-making.

The immature immune system of the neonates exaggerates the complex host inflammatory response in sepsis. The pro-inflammatory cytokines (IL-1β, IL-8, IL-6, TNF-α), acute-phase proteins (C-reactive protein, procalcitonin), and damage-associated molecular patterns (DAMPs) are released during the inflammatory cascade in neonatal sepsis.^7^ Upon infection, IL- 6 gets upregulated, preceding C-reactive protein synthesis, highlighting its clinical utility as an early diagnostic marker of infection and hyperinflammation. Serum CRP/high-sensitivity CRP are commonly used in laboratory settings but has limited use for further sub-classification in neonatal sepsis cases. Serum CRP, PCT and IL-6 levels have been studied extensively as biomarkers for identifying and differentiating sepsis cases from the healthy controls, but these individual biomarkers fail in further classifying the subtypes, most importantly the NS subgroup.^5^ Combining biomarkers as a signature always improves the diagnostic sensitivity in sepsis.^11^ Higher serum TNF-α levels indicate the onset of bacterial infection.^12^ Furthermore, IL-6 and TNF-α induce CCL20 production in various cell types, synergistically enhancing inflammatory responses. IL-6 and CCL20 are also involved in Th17 cell differentiation and recruitment. In contrast, anti-inflammatory cytokines such as IL-10 and TGF-β are key modulators of immune homeostasis during sepsis, limiting excessive pro-inflammatory responses. These cytokines act as immunological brakes, preventing the hyper-inflammatory state that could lead to tissue damage and organ dysfunction.^13^

In addition to cytokine dysregulation, alterations in serum micro- and macronutrients play a crucial role in the pathophysiology and progression of neonatal sepsis. Zinc deficiency, commonly observed in septic neonates, compromises immune function by affecting T cell development, cytokine production, and neutrophil activity. Similarly, selenium, an essential trace element with antioxidant properties, demonstrates notably reduced levels during septic episodes, potentially contributing to increased oxidative stress and cellular damage. Iron metabolism also undergoes significant alterations during sepsis, either deficiency or overload, potentially compromising immune response and increasing susceptibility to infection.^14^ Magnesium, an anti-inflammatory agent, serves as a cofactor for over 300 enzymatic reactions. Hypomagnesemia, along with hypocalcaemia, has been reported to cause Apparent Life- Threatening Events (ALTE) episodes and is positively correlated with sepsis severity.^15,16,17^ Conversely, elevated copper levels, especially in macrophages during infection, serve as an antimicrobial defence mechanism due to their toxic effects on the pathogens.^18^ Elemental profiling in neonatal sepsis may provide valuable insights into disease phenotype and progression, and offer potential supplementation strategies as additional therapeutic targets.

In this study, we comprehensively analysed serum inflammatory proteins and elemental composition in longitudinally followed-up neonates with sepsis at diagnosis and after treatment completion, and in healthy controls. Identified inflammatory markers were further validated in an independent cohort. A set of cytokines and trace elements was identified that could differentiate sepsis suspects from healthy controls and further differentiate between CP, CN, and NS cases. The identified deregulated elements have biological significance and could potentially be supplemented in addition to the treatment modalities in neonatal sepsis cases to improve clinical outcomes. The identified molecular signatures shed light on the sepsis disease mechanism and case presentation and hold potential for developing improved diagnostics to enable timely and accurate classification and initiate appropriate therapeutic interventions. On-time differentiation will equip clinicians to initiate targeted interventions before microbiological results are available, thereby improving patient outcomes in this vulnerable population, where time to appropriate treatment is crucial.

## Materials and methods

### Ethics committee approval

This multi-centre study was approved by the ethics committees of the Vardhman Mahavir Medical College and Safdarjung Hospital (IEC/VMMC/SJH/Project/2021-05/CC-157, dated 24.07.2021), All India Institute of Medical Sciences (IEC-1074/06.11.20, RP-28/2020), Dr. Baba Saheb Ambedkar Medical College and Hospital (5(32)/2020/BSAH/DNB/PF, dated 31.08.2020), Lady Hardinge Medical College (LHMC/IEC/2022/03/30), University College of Medical Sciences & Guru Teg Bahadur Hospital (IECHC-2022-51-R1, dated 07.12.2022), the International Centre for Genetic Engineering and Biotechnology (ICGEB/IEC/2021/28, version 2) and Jawaharlal Nehru University (JNU/IBSC/2022/82).

### Subject recruitment and classification

In this prospective cohort study, we enrolled preterm neonates born between 25 and 35 weeks of gestation who were admitted to participating hospitals’ neonatal intensive care units (NICUs) over 18 months after obtaining informed written consent from their legal guardians. Neonates with major congenital malformations or those born to mothers with HIV or hepatitis B, as well as those with postnatal age exceeding 28 days, were excluded from the study.

Neonates enrolled in the study were suspected of sepsis based on maternal and perinatal risk factors or clinical signs. Upon suspicion, a diagnostic work-up, including a sepsis screen and blood culture, was performed before initiating antibiotics. The sepsis screen assessed four parameters: low absolute neutrophil count (<1.5 × 10^9^ cells/L), elevated I/T ratio (>0.2), CRP (>6 mg/L), and/or erythrocyte sedimentation rate (ESR) >15 mm in the first hour, with any two abnormalities confirming a positive result. Neonates were classified into subgroups: culture-positive (CP) sepsis, culture-negative (CN) sepsis, no sepsis (NS), sepsis suspects (SS), recovered from sepsis (REC) and healthy controls (HC). CP cases had true pathogens in blood cultures, with coagulase-negative *Staphylococcus* classified as CP only if sepsis symptoms and antibiotic treatment for ≥ 5 days supported the diagnosis. Neonates were classified as CN sepsis if blood cultures were sterile or showed commensals, but sepsis screen results or clinical course indicated sepsis. NS cases did not meet CP or CN criteria. SS group consist of study subjects or cases that are suspected of sepsis but remain to be categorised. REC cases were CP or CN which recovered after treatment. Antibiotic decisions followed hospital-specific protocols. A set of neonates (n=44, CP/CN/NS/SS: 16/13/13/2) were followed up till recovery (details presented in Supplementary Table S1). Clinically stable neonates with no symptoms of sepsis within the first 5 days of life were enrolled as healthy controls (HC, n=12).

### Fractionation of whole blood for serum, plasma and CD45^+^ leucocytes

Whole blood was collected from the neonates at the time of diagnosis or follow-up, in BD Vacutainer® blood collection tubes, and an aliquot was loaded into the automated culture bottles (BD BACTEC or BACT/ALERT). The other aliquot was incubated at room temperature for 10-30 minutes, then centrifuged at 1,000 x g for 10 minutes to harvest the serum samples. These serum aliquots were transported and stored at -80 ^°^C until further processing. For plasma and blood cells, aliquots of independent blood samples were centrifuged at 2,000 × g for 10 minutes at room temperature and cell pellets were collected. The supernatant was centrifuged again at 4,000 × g for 15 minutes to completely deplete platelets. The plasma samples were treated with EDTA- Free Halt Protease Inhibitor Cocktail (Thermo Fisher Scientific, USA). The serum and plasma samples were transferred into pre-labelled cryovials and stored at -80 °C within 4 hours from blood collection. The cell pellet from blood samples was subjected to erythrocyte depletion using eBioscience™ RBC Lysis Buffer (Thermo Fisher Scientific, USA) according to the manufacturer’s instructions. The resulting leucocyte-enriched pellet was resuspended in cold MACS separation buffer (PBS, pH 7.2, 0.5% BSA, 2 mM EDTA, 10 °C). The CD45^+^ leucocytes were then isolated by positive magnetic selection using Dynabeads™ CD45 (Thermo Fisher Scientific, USA). The purified cells were immediately snap-frozen and stored at −80 °C until RNA extraction.

### Serum proteome profiling using Proximity Extension Assay for biosignature identification

Serum cytokines and chemokines of neonates with sepsis (n=38; CP/CN/NS: 13/13/12) at the time of suspicion and again after treatment completion and healthy controls (HC; n=12) were profiled using a Proximity Extension Assay (Olink® Target 96 Inflammation panel, Olink Proteomics AB, Salagatan 16F, SE-753 30 Uppsala, Sweden). Briefly, this technology utilizes a pair of antibodies labelled with unique DNA oligonucleotides, which bind to the target proteins. When these antibodies come into close proximity, their oligonucleotides hybridize, allowing DNA polymerase to extend the hybridized sequences and create unique PCR-amplifiable DNA reporter sequences for each detected protein. These reporter sequences are then amplified via PCR, quantified through high-throughput real-time PCR, and normalized using internal controls (Incubation control, Extension control and the Detection control) to generate Normalized Protein eXpression (NPX) values on a log_2_ scale. Higher NPX values suggest higher protein expression.

Detection panels undergo rigorous internal and external quality control (QC) procedures to ensure data accuracy and reliability. Internally, the QC process incorporates four distinct control measures: two immunoassay controls, one extension control, and one detection control. Externally, eight standardized samples are utilized to evaluate assay performance. These samples serve multiple critical functions, including the calculation of within-batch and between-batch coefficients of variation, the determination of the limit of detection, and facilitation of data normalization. Samples have passed QC when their NPX values deviate by less than 0.3 from the plate’s median NPX value. Scatterplots are generated to visualize the distribution of NPX values across all samples to identify potential outliers within the dataset, enabling the detection of any anomalies or deviations from expected patterns.

### GO enrichment analysis and pathway enrichment analysis

The deregulated protein sets were selected for GO and pathway enrichment analysis using ShinyGO 0.81 (https://bioinformatics.sdstate.edu;), and the top 20 GO terms and KEGG pathways were reported.^19^

### Serum micronutrient analysis using Inductively Coupled Plasma Mass Spectrometry (ICP-MS)

Due to limited sample availability, a subset of the study (CP/CN/HC: 5/7/7) was used for serum micronutrients (^57^Fe, ^66^Zn, ^63^Cu, ^77^Se, ^44^Ca, ^24^Mg) profiling using ICP-MS (iCAP-TQ mass spectrometer, Thermo Fisher Scientific, USA). In the next step, serum micronutrients were quantified in a separate validation set (CP/CN/NS/HC: 17/18/19/16). Briefly, serum samples (50 µL) were digested using 75 µL nitric acid (70%, #225711, Sigma, USA and 25 µL hydrogen peroxide (30%, #107298, Sigma-Aldrich, USA) in a microwave digester (Anton Paar, USA) at 140°C for 30 minutes. After cooling, the serum samples were diluted with ultrapure water (Honeywell, USA) to reduce the acid concentration to the permissible limit before being injected into the ICP-MS. The diluted acid-digested serum samples, along with the appropriate standard elemental mixture (#92091, Sigma-Aldrich, 0.1- 1000 ppm) were used for trace element quantification in the Kinetic Energy Discrimination (KED) mode, using helium as the carrier gas. The data were analyzed using Qtegra^®^ (Thermo Fisher Scientific) software, and the final concentration of the elements were calculated using the standard plot.

### Plasma Cytokine Quantification using Cytometric Bead Array (CBA)

The plasma cytokines TNF, IL-6, IL-8, and IL-10 were quantified using the Human Cytokine BD™ Cytometric Bead Array (CBA) using IL-8 (#51-9004049), IL-6 (#51-9004046), IL-1 (#51-9004060) and TNF (#51-9004040) beads, following manufacturer’s protocol (BD Biosciences, USA). Briefly, plasma sample (50 µL) was incubated with multi-analyte capture beads and phycoerythrin-conjugated detection antibodies. The beads were washed and data were acquired on a BD^®^ FACSLyric™ Flow Cytometer and analyzed using BD^®^ CBA analysis software. Absolute concentrations (pg/mL) for each analyte were extrapolated from an eight-point standard curve generated concurrently using protein standards present in the kit.

### Validation of IL-6 and IL-10 serum markers in an independent sample set

Serum levels of IL-6, and IL-10 were determined in an independent cohort of 80 samples (CP/CN/NS/HC- 20/20/20/20) using Enzyme Linked Immunosorbent Assays (ELISA MAX^TM^ Standard Set, BioLegend, USA) following the manufacturer’s instructions (Supplementary Table S1). Briefly, 96-well plates were coated with primary antibody in coating buffer. The plates were incubated at 4 °C overnight. Coated plates were washed three times with PBS-T (0.05% Tween- 20 in PBS). 200 µL of blocking solution [1% bovine serum albumin (BSA) in PBS] was added to each well, and the plates were incubated at room temperature (RT) for one hour. Serum of study subjects from the healthy control and patient groups were diluted 1:10, and 100 µL of the diluted samples was transferred to the ELISA plates after removing the blocking solution and washing the plates three times. After a 2 h incubation at RT, the plates were washed three times with PBS-T. 100 µL of detection antibody solution was added to each well, and the plates were incubated at RT for one hour. After washing, 100 µL of HRP substrate solution was added for 30 min. Then the plates are thoroughly washed 5 times with the wash buffer with a 1 min incubation per wash. 100 µL of TMB substrate was added to each well, and the plates were incubated in the dark for 15-30 minutes or until the desired colour developed. The enzymatic reaction was stopped by adding 100 µL of stop solution (1N Hydrochloric acid solution). The plates were read in a plate reader at 450 nm and 570 nm.

### qRT-PCR analysis of cytokines in neonatal leukocytes

The total RNA was isolated from CD45⁺ leucocytes using TRIzol™ Reagent (Thermo Fisher Scientific) and further purified using RNeasy® Plus Mini Kit (Qiagen, USA) to remove residual genomic DNA contamination. The RNA yield and purity were quantified using NanoDrop™ spectrophotometer (Thermo Fisher Scientific); only samples fulfilling purity thresholds (A_260_/A_280_ ≥ 2.0 and A_260_/A_230_ ≥ 1.8) were used for downstream applications. The cDNA was synthesized by using the SuperScript™ IV First-Strand Synthesis (Thermo Fisher Scientific), according to the manufacturer’s protocol. Quantitative real-time PCR (qRT-PCR) for TNFA, IL6, IFNG, IL1B, IL8, and IL10 was performed using the PowerTrack™ SYBR Green Master Mix (Thermo Fisher Scientific) on a QuantStudio™ 12K Flex Real-Time PCR System (Thermo Fisher Scientific) using primers described earlier.^20^ The thermal cycling conditions comprised initial activation at 95 °C for 60 seconds, 40 cycles of denaturation at 95 °C for 15 seconds, annealing/extension at 60 °C for 60 seconds, and a post-amplification melt-curve analysis to verify specificity. Relative gene expression was quantified using the 2^-ΔΔCt^ method, where target CT values were normalized to the geometric mean of glyceraldehyde-3-phosphate dehydrogenase (GAPDH) and beta-actin (ACTB) (ΔCT), and fold changes were calculated relative to the mean of healthy control (HC) samples (ΔΔCT), which served as the experimental calibrator.

### Statistical analysis

Statistical significance between groups was performed using univariate statistical tools like Welch’s t-test, parametric (unpaired or paired) and a p-value <0.05 was considered statistically significant. Proteins with a p-value <0.05 and log_2_fold change >±1.3 were selected as differentially expressed proteins (DEPs). Spearman (two-tailed) correlation analysis between the expression levels of DEPs, and trace elements was carried out using the GraphPad Prism version 8.4.2 for Windows (GraphPad Software, Boston, Massachusetts USA, www.graphpad.com). If the correlation value is close to ±1.0, it is considered the strongest correlation. Also, Spearman correlation coefficients (two-tailed) were calculated for IL6 and IFN-γ, and for IL-10 and IFN-γ. For targeted gene expression profiles (qRT-PCR) across clinical stages (HC, SS, CP+CN, and REC), statistical relevance was evaluated using a non-parametric Kruskal-Wallis test followed by Dunn’s post-hoc multiple comparisons test. As for plasma cytokine concentrations (CBA) across cohorts (HC, CP, and CN), data were transformed to a Log_10_ scale (pg/mL) and evaluated using one-way Analysis of Variance (ANOVA) followed by Dunnett’s multiple comparisons test against the HC baseline.

## Results

### Demographic details of the study subjects

The clinical details of the neonates (n=252, 32% female) belonging to sepsis (CP: n=67, CN: n=60), sepsis suspects (SS: n=10) and controls (no sepsis, NS: n=51; healthy control, HC: n=64) included in this study are presented in Supplementary Table S1. The study samples were randomly grouped using an online randomizer (www.randomizer.org). The discovery set comprised 88 samples from different sepsis sub groups (CP/CN: 13/13) and control (NS/HC: 12/12), of which all CP, CN, and NS cases were followed up till recovery, while the validation set comprised 20 samples from each group for ELISA, 28 for qRT-PCR, and 7 for CBA (CP/CN/HC/SS: 9/2/7/10). Micronutrient analysis was done in two cohorts; the discovery set consisted of 24 samples (CP/CN/HC: 8/7/9), and the validation set consisted of 70 samples (CP/CN/NS/HC: 17/18/19/16).

The summarised clinical characteristics of neonates enrolled in both the discovery and validation studies are presented in a heat map (Figure 1). Females accounted for 40% of each study group in the discovery set, while they accounted for 29% of the total samples used in the validation set. *Acinetobacter baumannii*, *Escherichia coli*, *Klebsiella pneumonia* and coagulase-negative Staphylococcus (CoNS) were isolated in neonates with CP sepsis. In the discovery set, 7.0% and 23.0% of CP cases were contributed by *Acinetobacter baumannii* and *Klebsiella pneumonia*, respectively. The prevalence of *Acinetobacter baumannii* and *Klebsiella pneumonia* in the CP groups of the independent validation set was 10% and 10%, respectively. Most CP and CN sepsis cases in neonates were early-onset sepsis (EOS). The gestational age of neonates in the CN (30.2±1.6 weeks) sepsis group was significantly lower than that of the HC group (32.4±1.0 weeks, p<0.0005) and the NS groups (32.4±1.4 weeks, p<0.002) in the discovery set, but was not significant for the CP group (32.5 ±1.5 weeks). The gestational age was also significantly lower in the sepsis groups (CP/CN: 30.9±1.9/31.05±1.7 weeks) compared to HC (33.35±1.0 weeks, p<0.008) but was insignificant with NS (31.7±1.7 weeks) in the validation set. The birth weight of the neonates of the discovery set was lower in CN (1.3±0.2 kg) compared to the HC (1.6±0.3 kg, p<0.007) and NS (1.6±0.4 kg, p<0.05) groups. Similarly, lower birth weight was observed in the validation set for CP (1.5±0.4 kg) and CN (1.5±0.3 kg) compared with HC (1.92±0.3 kg; p<0.007). In the discovery set, the cesarean delivery rate was lower in the CN group (38.5%) and higher in the CP group (61.5%) as compared to HC (54.5%) and NS (58.3%). On the contrary, in the validation set, the number of cesarean deliveries was lower in the CP group (35%) and higher in the CN group (50%) than in the HC group (40%). In the discovery set, the absolute neutrophil count was lower in the CP group (3.7±1.4×103) compared to CN (9.9±10.5×103, p<0.05) and NS (6.3±2.5×103, p<0.004) groups. In the validation set, the absolute neutrophil count was lower in the CP group (2.8±2.7×103) compared to the NS (5.4±2.4×103, p<0.02) groups.

**Figure 1:**
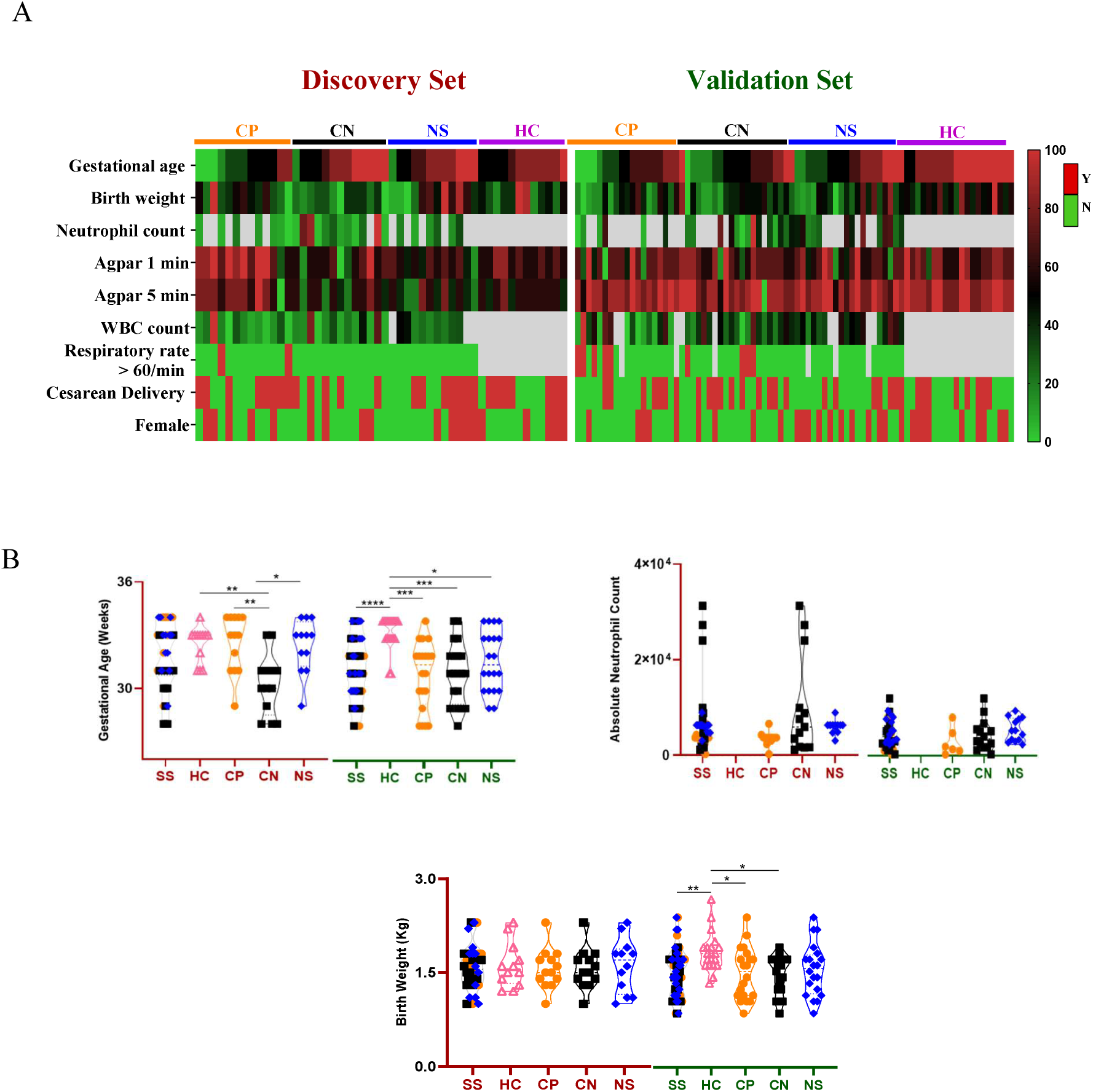
Epidemiological details of the study subjects and groups used in this study. **A.** Clinical characteristics (gestational age, birth weight, caesarean deliveries, absolute neutrophil count) of the study participants used in the discovery and validation set (Y/N:Yes/No) (B). Statistical comparisons between groups (P-value<0.05:*, <0.005:**, <0.0005:***, <0.0001:****). The details of the study subjects are presented in Supplementary Table S1.

### The serum inflammation protein signature showed group-specific variations in neonatal sepsis

Of the 92 proteins (details in Supplementary Table S2) monitored across all study groups, 89 passed QC (as described in the method section). Three proteins (β-NGF, IL-24, and Thymic stromal lymphopoietin (TSLP)) had a coefficient of variation (CV) of more than 25 % of CV and were excluded from the study. A few serum samples (CP, n=4; CN, n=2; NS, n=3; HC, n=1) did not pass the QC (as described in the method section) and were excluded. Between CP and HC, 7 DEPs (log_2_FC > ±0.4, p-value < 0.05) were identified (Figure 2B). And 3 DEPs (log_2_FC > ±0.4, p-value < 0.05) were identified between CN and HC (Figure 2B). Compared with HC, the CP group showed significantly elevated serum levels of IL-10, IL-6, LIF, and TNF-α, and lower levels of CCL28 and IFN-γ (Figure 2B). Serum IFN-γ, CXCL10, and CCL11 levels were significantly lower in the CN group (Figure 2B). Between CP and CN, a significantly higher abundance (log_2_FC> ±0.4, p-value < 0.05) of TNF-α, IL-10, IL-6, CCL20 and MCP-3 was observed (Figure 2B). Between the CP and NS groups, TNF-α, MCP-3 and CCL20 qualify the criteria (log_2_FC> ±0.4, p-value < 0.05) (Supplementary Figure S1A). Serum IL-1α levels in the CN groups were significantly high (log_2_FC> ±0.4, p-value < 0.05) compared to the NS groups (Supplementary Figure S1B). A set of 3 DEPs (FGF-23, IFN-γ, and CCL11) (log_2_FC > ±0.4, p-value < 0.05) was identified between the NS and HC groups. Serum FGF-23 levels in the NS group were significantly high, and IFN-γ and CCL11 were low (Supplementary Figure S1C). Out of these 12 DEPs, at least between two groups, a set of five molecules (IL-6, LIF, IFN-γ, CCL11, FGF-23) showed significant deregulation, including between sepsis suspects and HC (Figure 2C).

**Figure 2.**
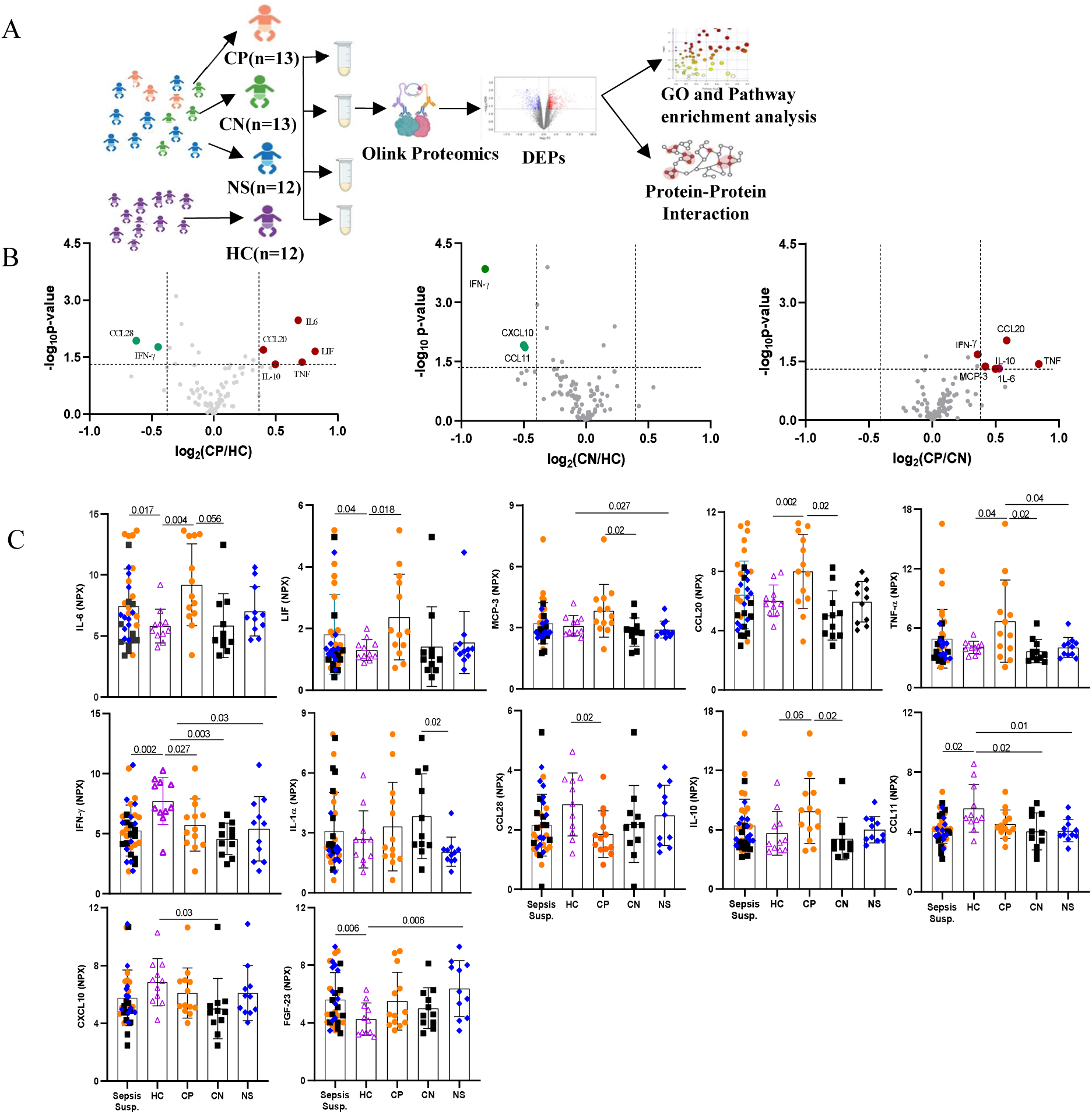
Serum inflammatory proteins profiled using Proximity Extension Assay (PEA) showed differential expression in neonatal sepsis cases compared to healthy controls (HC). **A.** Schematic diagram representing the workflow used for Olink Proteomics analysis. **B.** Volcano plots showing dysregulated proteins (log_2_FC>±0.4, p-value < 0.05) between study groups: Culture Positive (CP) and Healthy control (HC), Culture Negative (CN) and HC, CP and CN. **C.** Box plots showing the abundances of dysregulated analytes between Sepsis suspects and Healthy controls (HC) and their group-specific changes between CP, CN and NS.

### Serum micronutrient levels in neonatal sepsis patients showed variations in trace element homeostasis

The ICP-MS analysis determined discrete differences in the serum concentrations of all six trace elements (^57^Fe, ^66^Zn, ^63^Cu, ^77^Se, ^44^Ca and ^24^Mg) in the sepsis cohorts. Among these, serum iron (^57^Fe) concentrations were significantly decreased (p<0.05) in both CP and CN sepsis subjects compared to HC (Figure 3A). A similar profile was determined in both serum selenium and zinc in both CP and CN cohorts exhibiting lower levels than HC (Figure 3B). Additionally, copper (^63^Cu) levels in the CN groups were significantly (p<0.05) lower than in the HC (Figure 3B). The remaining elements were similar across the study groups. In the recovered CP subjects, selenium levels recovered, while serum iron levels were similar to the initial levels observed at the time of diagnosis. The serum Fe and Cu levels in the CN groups completing therapy recovered to normal levels (Figure 3C). In a separate validation set (CP/CN/NS/HC: 17/18/19/16), ^77^Se, ^24^Mg, and ^44^Ca showed a significant difference between the sepsis subsets and the healthy control (Supplementary Figure S2). Serum selenium levels in the CP were significantly lower compared to the other groups, as previously seen (Supplementary Figure S2). Additionally, magnesium and zinc were lower in the CP study subjects than in the CN subjects. Iron levels showed a decreasing trend in CP compared to HC (Supplementary Figure S2). CP study subjects also had lower calcium levels than the NS study subjects. Compared with HC, magnesium and calcium levels were higher in both CN and NS (Supplementary Figure S2).

**Figure 3:**
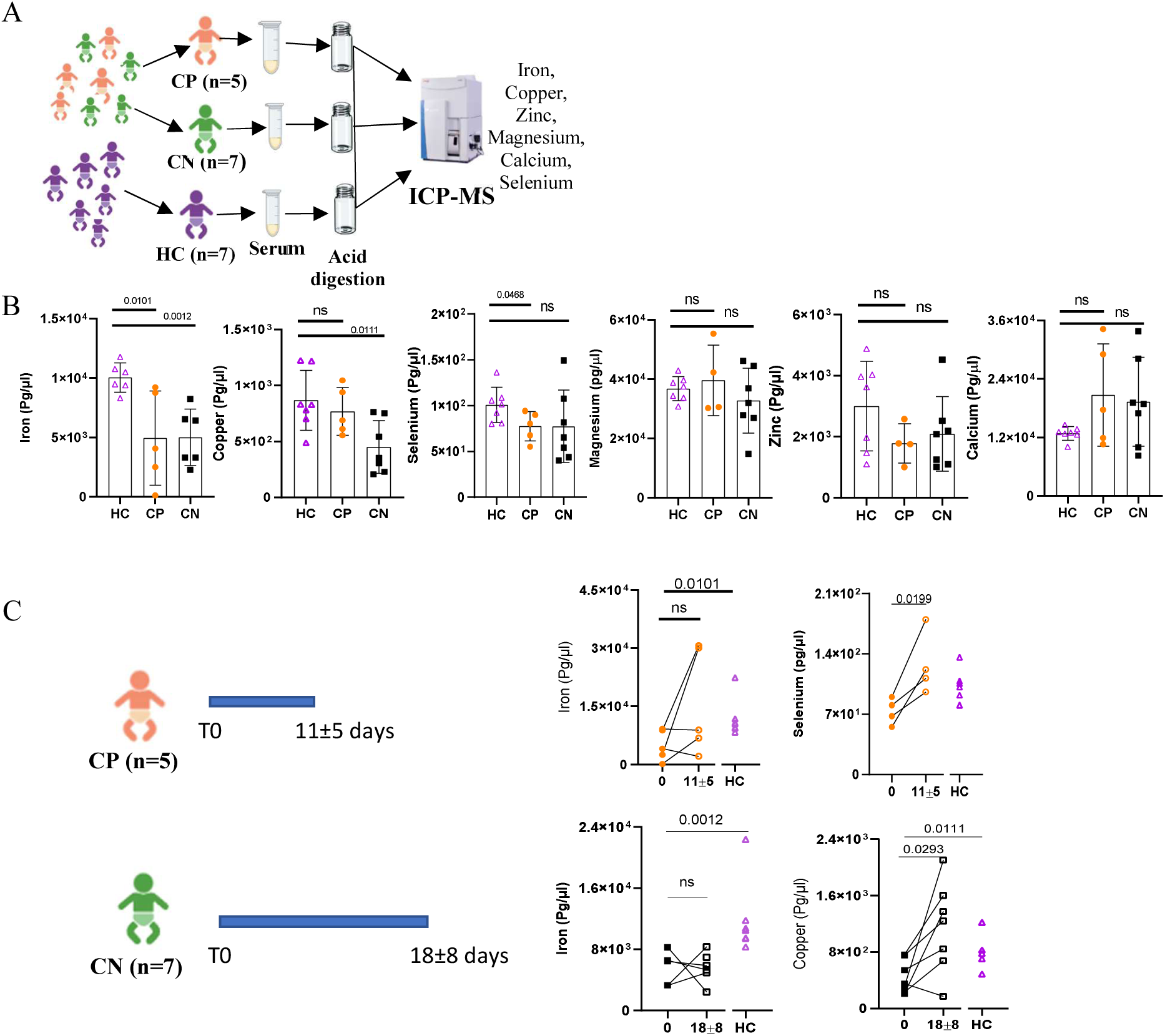
Serum elemental composition of sepsis patients showed variations. **A.** Schematic diagram representing the workflow used for ICP-MS. **B.** Box plot of serum Iron, Copper, Calcium, Magnesium, Zinc, and Selenium between CP, CN and HC. **C.** Box plots depicting the serum Iron, Selenium, and Copper levels in follow-up neonatal sepsis cases upon recovery compared to HC.

### Protein-protein interaction analysis of the deregulated serum proteins and elements showed dysregulation of the TNF, JAK-STAT, and NF-Kappa B pathways between sepsis subgroups

The DEPs identified from the proteomics experiments were selected for protein- protein interaction (PPI) analysis using the Cytoscape tool (Figure 4A). The identified perturbed pathways with FDR < 0.05 between CP and HC included TNF, JAK-STAT, and T cell receptor signalling pathways (Figure 4B). Further in the CN, the IL-17, Th17, TNF and chemokine signalling pathways were dysregulated (Figure 4B). Between the CP and CN groups, the TLR, chemokine, IL-17, TNF, and NF-Kappa B signalling pathways were perturbed (Figure 4B). A positive correlation between IL-6 and IFN-γ was observed only in the CP group (Figure 4C). Between serum IL-6 and IL-10, a significant positive correlation was observed in the CP and CN sepsis groups (Figure 4C). A significantly higher IL-6/ IFN-γ ratio was observed in the sepsis suspect groups (CP, CN and NS) compared to HC (Figure 4D).

**Figure 4:**
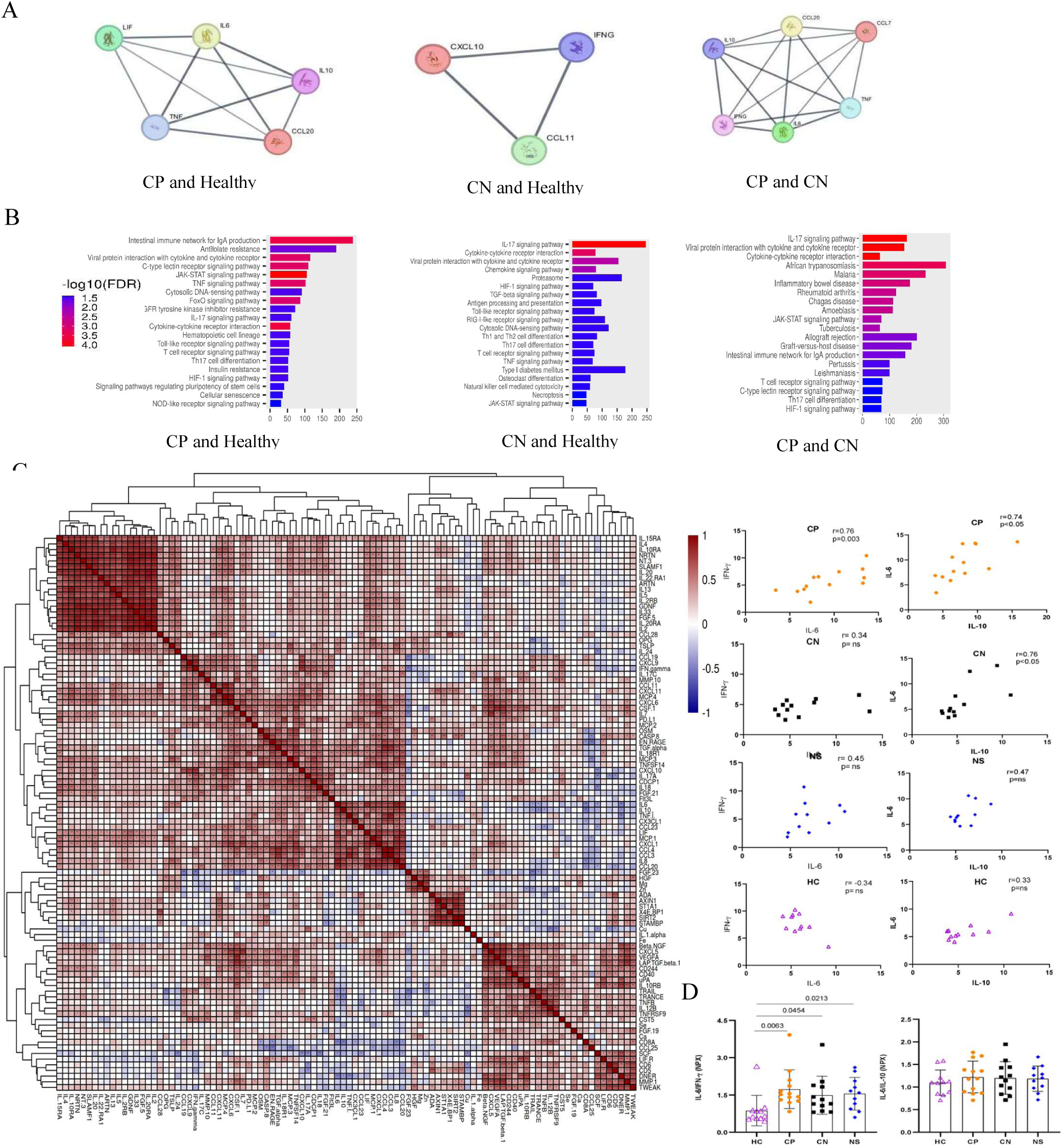
Protein-Protein interaction, pathway analysis and correlation analysis between sepsis and control groups. **A.** The protein-protein interaction network of the sorted DEPs across the different subgroups of neonatal sepsis was constructed using Cytoscape (confidence score ≥0.7). **B.** Bar plot depicting the significantly dysregulated pathways between the sub- groups. **C.** Correlation heat map (Pearson r values) of inflammatory proteins and microelements. Red indicates a positive correlation, and blue shows a negative correlation. Correlation (Spearman r) between IL-6 and IFN-γ, IL-6 and IL-10. **D.** Box plots with IL-6/IFN- γ ratios and IL-6/IL-10 ratios between the sub-groups.

### Serum inflammation markers in neonatal sepsis cases receiving treatment show specific variations

The sepsis neonates were followed up until they completed antibiotic therapy and recovered from clinical symptoms. IL-6 recovered and reached the control (HC) level, while the rest remained similar (Figure 5A). Upon completion of treatment, the serum IFN-γ, CXCL10, and CCL11 levels in the CN groups showed significant variation and were closer to those of the HC group (Figure 5B). In recovered NS patients, serum FGF-23 and CCL11 levels showed significant changes (p<0.05), returning to levels similar to those of HC, whereas the other two dysregulated proteins remained unchanged. (Figure 5C).

**Figure 5:**
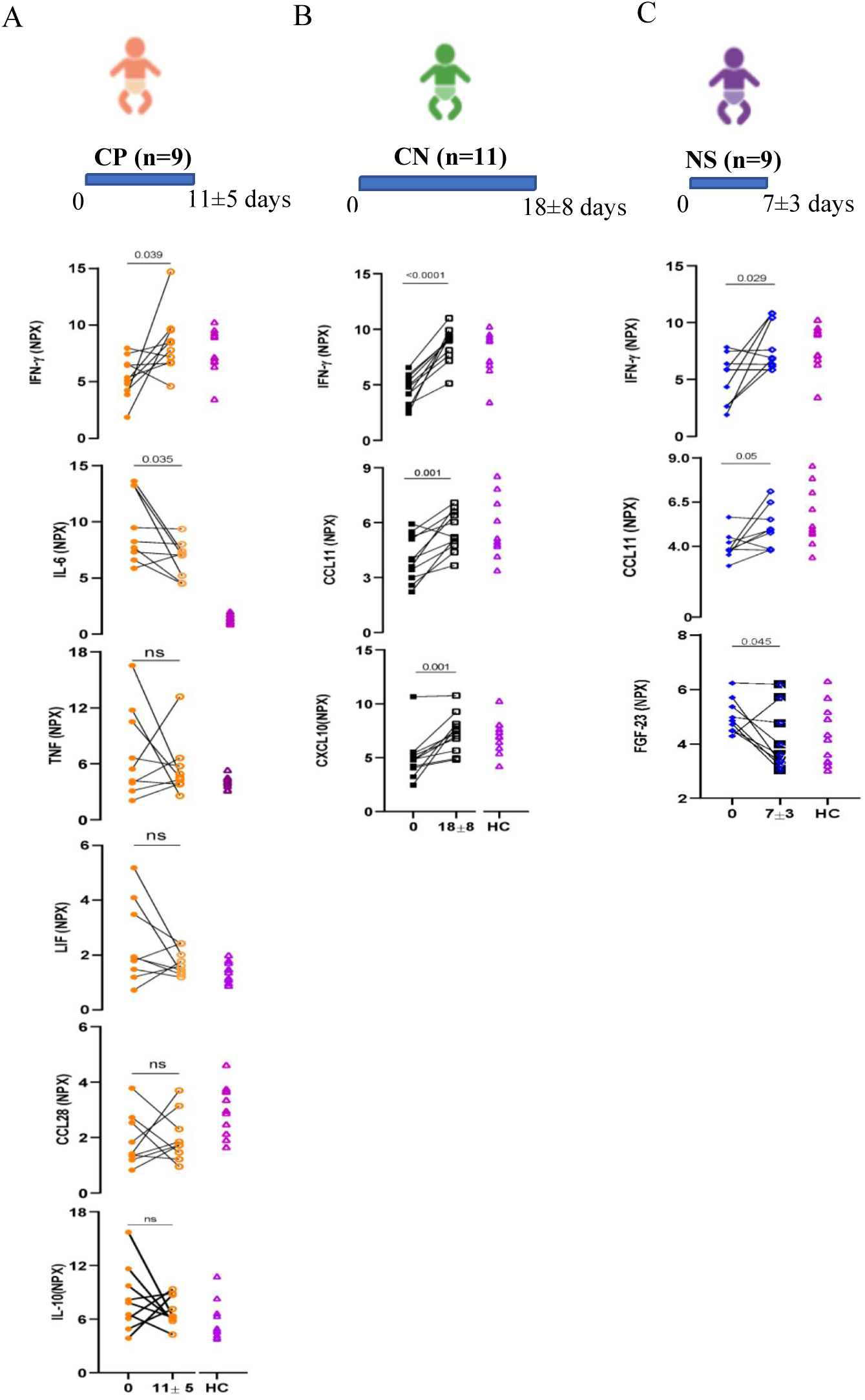
Box plots showing the trend of important serum proteins in the follow-up culture-positive, culture-negative, and no-sepsis groups. Abundance of important serum protein at diagnosis and recovery in culture positive (**A**: CP), culture negative (**B**: CN) and no sepsis groups (**C**: NS).

### The mRNA expression of cytokines in CD45+ leucocytes showed variations in sepsis cohorts

The mRNA expression profiles of *TNFA*, *IL6*, *IFNG*, *IL1B*, *IL8*, and *IL10* in circulating CD45^+^ leucocytes from the four neonatal cohorts exhibited discrete shifts in transcript abundance across sepsis progression. Sepsis suspicion (SS) cohort exhibited upregulation of *IL6* (2.56 ± 0.80-fold), *TNFA* (5.45 ± 2.17-fold), and *IL8* (3.14 ± 1.05-fold), (p ≤ 0.01) compared to healthy control (HC), prior to confirmation of sepsis (Figure 6A, B, E). The clinically confirmed sepsis; either culture positive (CP) or culture negative (CN), exhibited more robust upregulation of these cytokines including the *IL6* (11.66 ± 3.49-fold; p < 0.0001), *TNFA* (4.61 ± 1.45-fold; p < 0.0001), and *IL8* (5.38 ± 2.28-fold; p < 0.0001) compared to healthy control (HC) (Figure 6A, B, E). Furthermore, *IL1B* (2.84 ± 1.08-fold; p < 0.0001) exhibited robust upregulation, whereas *IFNG* (1.93 ± 0.54-fold; p = 0.003) was marginally upregulated in this cohort (Figure 6C, D). The anti-inflammatory cytokine *IL10* showed a stepwise modulation, from 2.72 ± 0.99-fold in the SS cohort (p = 0.03) to 4.35 ± 0.77-fold in the CP+CN cohort (p < 0.0001) compared to healthy control (HC) (Figure 6F), reflecting a compensatory response to the surging pro-inflammatory profiles. However, in other independent samples, *IL10* exhibited a significant variation, with high expression in this cohort (data not shown). In general, the post-recovery (REC) cohort exhibited lower relative expression of *IL6* (2.45 ± 0.64-fold; p = 0.0006), *TNFA* (1.94 ± 0.58-fold; p = 0.01), *IFNG* (1.19 ± 0.13-fold; p = 0.03) and *IL8* (2.35 ± 0.91-fold; p = 0.04) compared to sepsis positive cohort reflecting the clinical improvement (Figure 6A, B, C, E).

**Figure 6.**
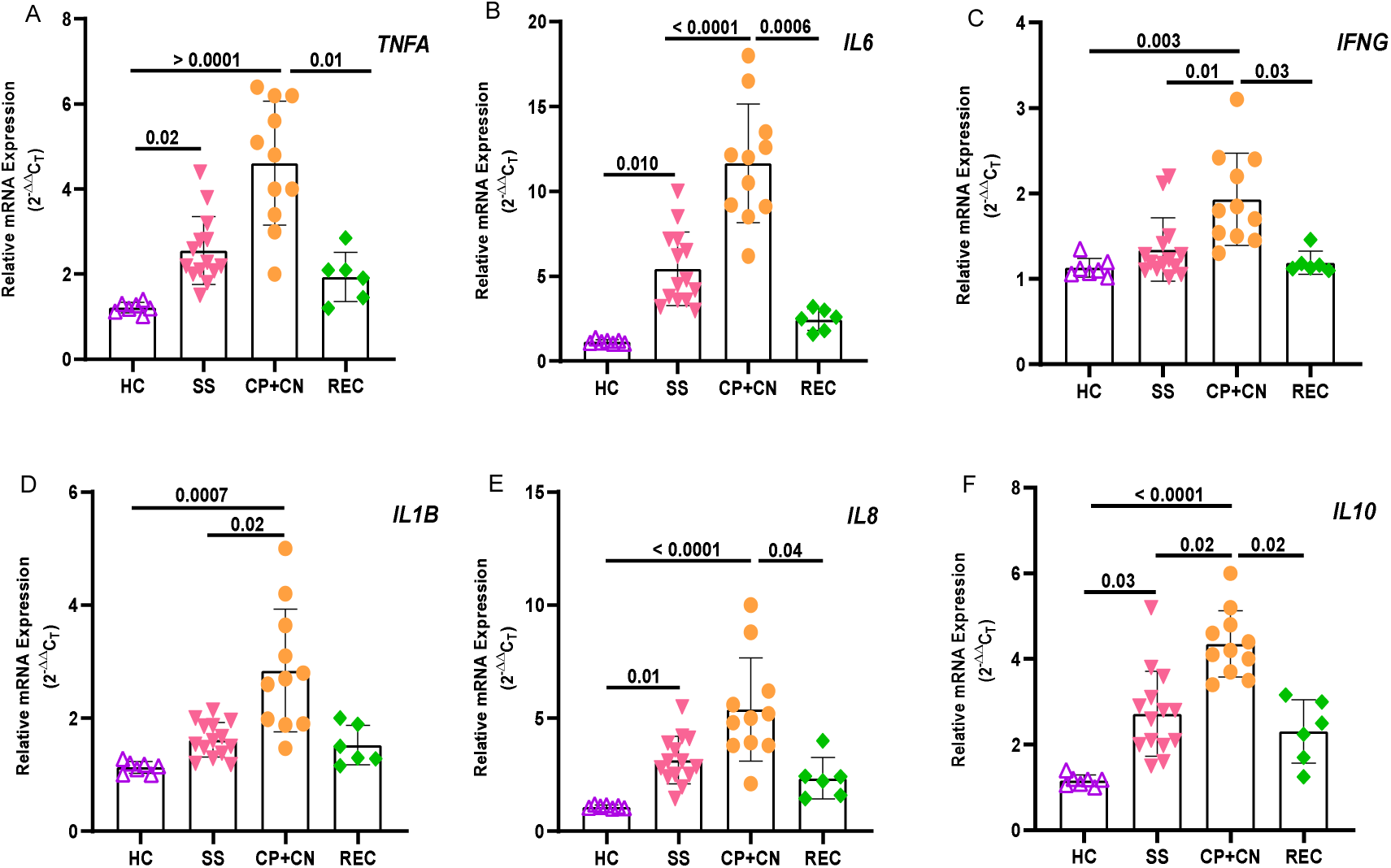
Cytokine mRNA expression in circulating CD45^+^ leucocytes across clinical stages of neonatal sepsis. Relative mRNA expression of (A) *TNFA*, (B) *IL6*, (C) *IFNG,* (D) *IL1B*, (E) *IL8*, and (F) *IL10* in CD45^+^ cells isolated from whole blood. Cohorts are defined as healthy controls (HC), sepsis suspicion (SS), confirmed sepsis (CP+CN; culture-positive and culture-negative sepsis combined), and post-recovery (REC). Target transcripts were quantified by qRT-PCR and normalized to the geometric mean of two endogenous controls *(ACTB* and *GAPDH*). Fold change was calculated using method 2^-ΔΔC^ relative to the mean of the HC cohort. Data points represent the mean of technical replicates for individual neonate samples; bars indicate cohort mean ± SD. p-values indicated above the comparison brackets were determined by a Kruskal-Wallis test followed by Dunn’s multiple comparisons test.

**Figure 7:**
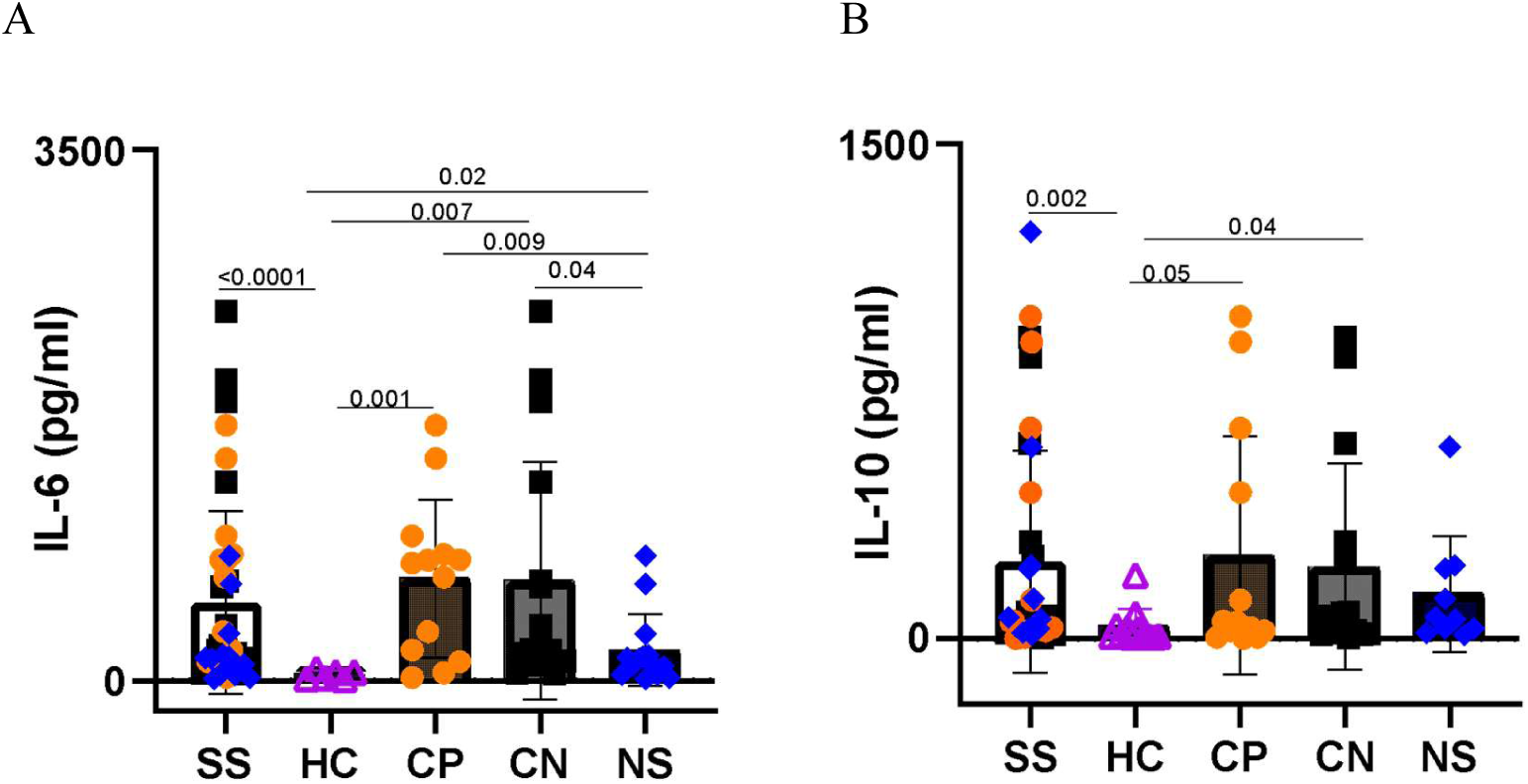
Neonatal serum IL-6 and IL-10 levels, in an independent validation set (n=80) showed sepsis-specific trends: Box plots showing IL-6 **(A)** and IL-10 **(B)** levels in the serum of validation culture-positive (CP), culture-negative (CN), and no-sepsis (NS), and healthy control (HC) groups. Sepsis suspects (SS) include all CP, CN and NS subjects used in this study.

**Figure 8:**
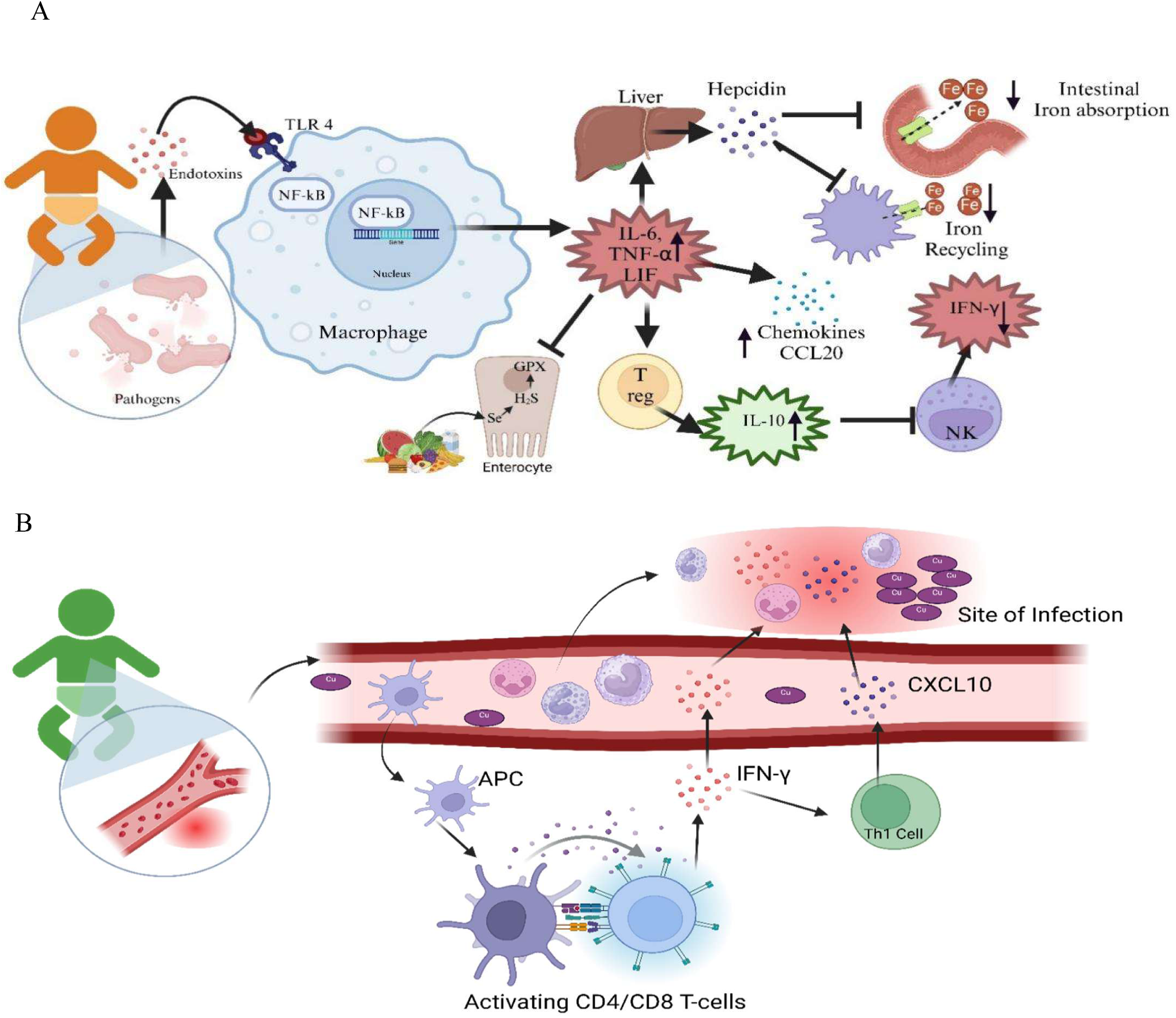
Schematics representing the pathophysiological changes in **A**. culture-positive (CP) and **B.** culture-negative (CN) neonatal sepsis cases.

### Validation of Serum Il-6 and Il-10 levels in the serum or plasma of an independent cohort

In sepsis-suspected neonates, serum IL-6 concentrations were significantly elevated (p<0.0001) compared to HC (293.11 ± 678.43 pg/ml). All sepsis subgroups exhibited significantly higher IL-6 levels relative to HC: CP sepsis (1337.73 ± 1827 pg/ml), CN sepsis (880 ± 1194.7 pg/ml), and NS (345.6 ± 605.99 pg/ml). Furthermore, both the CP and CN groups exhibited elevated IL-6 concentrations compared to the NS group.

Similarly, IL-10 levels were significantly increased (p<0.002) in sepsis-suspected neonates compared to HC (36.0 ± 53.5 pg/ml). The CP group showed markedly elevated IL-10 concentrations (838.63 ± 1304.5 pg/ml), while the CN group also demonstrated significantly higher IL-10 levels (381.43 ± 717.10 pg/ml) than the HC group. The NS group (236.09 ± 370.50 pg/ml) showed lower IL-10 levels than CP and CN, but the differences were insignificant. Multiplex cytokine assays (IL-6, TNF, IL-8, and IL-10) in plasma also revealed similar changes in the CP, CN, and HC groups (Supplementary Figure S3).

## Discussion

Neonatal sepsis represents a potentially devastating systemic infection, which can be mono or polymicrobial in origin, and it poses a significant threat to newborns due to their immature immune systems and nonspecific clinical presentations. This condition remains a leading cause of neonatal mortality worldwide, particularly in developing and underdeveloped countries, emphasizing the critical importance of early recognition and prompt therapeutic intervention.^21^ Based on the blood culture and mass spectrometry results, neonatal sepsis patients were grouped into CP, CN and NS. These diagnostic methods take 24-48 hours to yield results, exposing neonates to broad-spectrum antibiotics during the early phase of their lives. Timely classification of the neonates using appropriate immune biosignatures would help to initiate treatment modalities to improve clinical outcomes. This study identified a set of inflammatory markers and a micronutrient profile that would help distinguish between sepsis subgroups (CP, CN, and NS). This identification of a biosignature would help with faster diagnosis and more accurate treatment, enabling targeted therapies.

In this study, the CP sepsis group showed a distinct inflammatory signature characterized by elevated levels of pro-inflammatory cytokines IL-6, IL-10, LIF, and TNF-α, decreased IFN-γ, and CCL28 levels. Extensive immunological studies focusing on IL-6, LIF, OSM, and IL-27, all belonging to the IL-6 cytokine family, have highlighted their key roles in host defence mechanisms, demonstrating pivotal contributions to antimicrobial and antiviral immunity by providing essential tissue protection against infection-induced pathological injury.^22,23,24^ Recent studies have also reported IL-6 levels to be high in neonates with sepsis and their levels significantly reduced upon treatment.^25^ Accordingly, higher serum IL-6 and LIF levels observed in the CP group compared to the HC indicate an active immune response against infection, and upon recovery, IL-6 levels are similar to those of the HC.

Among immune cells, monocytes and macrophages are the primary sources of TNF-α.^26^ TNF- α, along with IL-1, is one of the few inflammatory mediators often reported at inflammation sites during the initial phase.^27^ In neonatal sepsis, TNF-α causes increased vasodilation and vascular permeability, leading to systemic edema and, ultimately, septic shock. Especially in early-onset neonatal cases, these pathological changes can rapidly progress to multiple-organ failure and death.^28^ Consistent with these findings, we also observed that the CP sepsis groups exhibited a similar trend, characterized by elevated TNF-α levels at the time of suspicion.

IFN-γ is mainly released by activated CD4+ and CD8+ T cells and NK cells, enhancing HLA-DR expression and antigen presentation, thereby enhancing immune response capabilities.^29^ Both pre-clinical and clinical sepsis studies show a significant reduction in T cell IFN-γ production, and therapeutic interventions to restore this cytokine improve survival in experimental models.^30^ The critical immunoregulatory role of IFN-γ is further emphasized by observations in infants with deficient production, characterized by compromised neutrophil mobility, reduced NK cell activity, and an increased susceptibility to recurrent infections.^31^ Furthermore, patients with severe sepsis exhibited reduced IFN-γ production and an imbalance in T helper cell cytokines, characterized by elevated Th2 cytokines and decreased Th1 cytokines.^32^ In this study, similar observations in both CP and CN sepsis groups were witnessed, marked by lower IFN-γ level at diagnosis, which strongly suggests a dysregulated immune response contributing to sepsis – associated immunosuppression. The IFN-γ levels are partially resolved upon treatment completion in recovered neonates, indicating an improvement in the immune response. An elevated IL-6/IFN-γ ratio, as observed in infectious diseases, may drive exuberant inflammation and subsequent tissue damage.^33^ In this study, a positive correlation (r=0.77, p<0.05) between IL-6 and IFN-γ was observed in CP sepsis, and a significant increase in the IL-6/IFN-γ ratio was observed in the sepsis suspects than in HC.

Various immune cells, encompassing T and B lymphocytes, monocytes, NK cells and macrophages, synthesize and release IL-10.^34^ This cytokine suppresses the production of pro-inflammatory mediators, including IL-1, IL-6, GM-CSF and TNF-α in cells of the immune system.^35^ In this study, we observed significantly elevated IL-10 concentrations in the CP group, which corresponded with decreased levels of IFN-γ. Interestingly, levels of other pro-inflammatory cytokines, such as IL-6 and TNF-α, remain higher in the CP group, as the majority of cases in our study group are early-onset sepsis (EOS), in which the pro-inflammatory effect predominates over the anti-inflammatory effect. Studies have revealed a positive monotonic relationship between IL-6 and IL-10 in sepsis patients infected with Gram-positive and Gram-negative bacteria. While this correlation was significant, it was particularly pronounced in sepsis patients infected with Gram-negative bacteria, demonstrating a stronger linear association between these two cytokines.^36^ In our study, most of the CP patients were reported with Gram-negative pathogens, and a significant positive correlation between IL-6 and IL-10 was observed (r=0.75, p<0.05). Interestingly, a positive correlation between serum IL-6 and IL-10 levels was also observed in the CN patients (r=0.77, p<0.05). CCL20 is an important chemokine that plays a crucial role in inflammatory responses, as it mainly recruits CCR6- expressing cells, including memory T cells, B cells, Th17 cells, and immature dendritic cells.^37^ In chronic inflammation, IL-6 and CCL20 form a positive feedback loop where IL-6 induces CCL20 production, which recruits more inflammatory cells that produce IL-6.^38^ TNF-α can also upregulate CCL20 through NF-κB signalling pathways.^39^ High serum CCL20 levels observed in the CP patients corroborate earlier reports.^25^

IFN-γ-inducible protein 10 (IP-10/CXCL10), a pivotal chemokine secreted by cells stimulated with type I and II interferons and lipopolysaccharide, acts as an important chemoattractant for activated T lymphocytes.^40^ This molecular mediator demonstrates prominent expression in Th1-mediated inflammatory pathologies, playing a fundamental role in orchestrating activated T cell recruitment to inflammatory tissue microenvironments.^41^ This complex immunological relationship between IFN-γ and CXCL10 is also observed in our data by the robust linear relation, wherein diminished IFN-γ levels directly corresponded to decreased CXCL10 concentrations, and subsequent immunological recovery is marked by a simultaneous elevation of both cytokine and chemokine levels. Interestingly, low serum levels of IFN-γ and CXCL10 were reported in neonates with inflammatory diseases, such as placental histological chorioamnionitis (HCA), using Olink analysis.^42^ Overall, the downregulation of CXCL10 and IFN-γ shows overall repressed Th1 immunity in neonatal sepsis. These important molecules, when combined as a biosignature, could distinguish between the CP, CN and NS sub-groups of neonatal sepsis cases.

The serum levels of the inflammatory markers TNF-α, MCP3, and CCL20 were higher in CP than in NS, and IL-1α levels were higher in CN than in NS. This signature can not only distinguish between CP and CN but also from NS controls, facilitating early diagnosis and helping initiate appropriate therapeutic interventions. Inflammatory signature consisting of IL- 6, IL-10, TNF-α, MCP3 and CCL20 was higher in CP in comparison to CN. Based on our findings, the identified biosignature appears to better classify sub-classes of neonatal sepsis cases for which limited solutions are available and demonstrates the importance of using biosignatures rather than using single markers like IL-6 or IL-10.

We also observed that iron levels were much lower in sepsis-confirmed cases than in HC. This dysregulation aligns with our observation of increased IL-6 levels in sepsis, which is known to modulate iron homeostasis.^43^ IL-6 can stimulate hepatocytes to secrete various acute-phase proteins, including hepcidin (an iron regulatory hormone), CRP, fibrinogen, complement C3, thrombopoietin, and ferritin.^44^ Briefly, during inflammation, the IL-6 level increases, which induces the release of hepcidin from hepatocytes through the JAK/STAT3 pathway. The released hepcidin binds to the iron exporter, ferroportin and causes its internalization, ubiquitination and lysosomal degradation. As a result, intracellular iron concentration increases significantly, and to sequester this iron, ferritin levels increase in the cells, leading to decreased serum iron levels, as observed in the CP and CN sepsis groups.^45, 46^ The validation set with a larger cohort showed similar trends with lower iron levels in CP.

Selenium (Se) is an essential trace element in many biological processes, mainly by regulating selenoprotein synthesis, an important class of protein with antioxidant and anti-inflammatory properties.^47^ Se deficiency is often observed in infection and is also reported in sepsis-affected neonates. Se deficiency greatly impairs innate and adaptive immunity by reducing NK cell activity, T cell function and immunoglobulins (IgM and IgG) while increasing reactive oxygen species (ROS). This compromised immune function, combined with diminished neutrophil and macrophage activities, increases susceptibility to infections.^48^ Elevated serum IL-6 and TNF-α levels observed in the sepsis-suspected cases, control the Se levels by influencing its bioavailability. Adequate Se levels are essential for promoting the expression of selenoproteins and IFN-γ, which could contribute to low IFN-γ levels in sepsis-suspected cases.^49^ Our study also showed that serum selenium levels were low at suspicion, which, upon recovery, returned to normal levels, indicating the antioxidant attributes of selenium and its possible influence in diminishing mortality related to sepsis in neonates. In the separate validation cohort, we also observed selenium deficiency in the CP group compared with other sepsis subsets and the healthy control. This also highlights the need for further research on how sepsis affects selenium levels and the potential of selenium supplementation as a therapeutic approach to manage neonatal sepsis. Copper accumulation observed during fungal infections is primarily driven by the host immune system as an antimicrobial strategy. When copper levels are elevated by the host, it creates oxidative stress conditions around the pathogen; it can disrupt cell membranes, interfere with essential proteins and enzymes and lead to the generation of harmful ROS.^50^ This clearly explains the low copper levels observed in the CN group’s serum. And among subjects completing therapeutic intervention, there was also a significant increase in serum copper levels, reaching healthy levels.

Zinc plays a major role in immune activation and is required for a wide range of functions, including transcription, translation, and maintaining homeostasis.^51^ Zinc deficiency leads to Th1 and Th2 imbalance, resulting in reduced IFN-γ production.^52^ This decrease is seen in the CP group, correlating to the serum zinc levels in the culture-positive group of the validation set. Low and high serum calcium levels were correlated with increased mortality.^53^ The serum calcium levels in sepsis patients showed fluctuations during hospitalisation, therefore it is challenging to categorize patients based on baseline calcium levels. Tracking calcium levels longitudinally would help understand disease progression and enable timely interventions.^54^ We observed that serum calcium levels were higher in the sepsis group in comparison to the healthy control in the validation set. Even though neonatal sepsis is correlated with hypomagnesemia, hypermagnesemia is related to increased inpatient mortality.^55^ This is mainly attributed to the role of magnesium as a calcium channel blocker, resulting in disruption of cardiac function and heart failure, and also the effect of magnesium on blood pressure and transmission of impulses.^56,57,58^ This is observed in the validation set, with higher serum magnesium levels in sepsis groups compared to healthy controls.

The levels of ^57^Fe, ^66^Zn, ^63^Cu, ^77^Se, ^44^Ca, and ^24^Mg were lower in CP than in the other sepsis subgroups, indicating the important role of these elements in the pathogenesis of culture- positive sepsis. The serum elemental composition can distinguish between the sepsis subtypes. This shows that serum elements can be used as targeted therapeutics to improve the immune response in neonatal sepsis. Further research is needed to understand the mechanism of action and the potential for supplementation.

Although neonatal PBMCs are not programmed to be highly responsive to PAMPs, we observed significant levels of Th1 cytokines and a well-defined immunometabolic network even in early stages (suspected EOS) of sepsis. Besides circulating PBMCs, cytokines are also produced by endothelial cells in the neonates and their mechanisms of immune dysregulation and pathobiology are distinct from those of PBMCs.^59^ In order to further confirm the source of the cytokines and related immune determinants determined by Olink and immunoassays, we investigated the mRNA expression of key cytokines in circulating PBMCs from venous blood. Since endothelial cells are also present in neonatal blood, we employed CD45^+^ sorting to specifically select PBMCs (endothelial cells are CD45^-^). These mRNA expression profiles exhibited similar cohort-specific modulation of major cytokines, including IL-6, IL-8, and TNF, underscoring the role of PMBCs in soluble cytokine production in neonatal sepsis. Further research is needed to understand the reasons for some mismatch observed between the mRNA expression profiles of some cytokines (IFN-γ and IL-1β) and their protein profiles in sepsis.

Olink-based cytokine profiling requires minimal sample volume and provides high throughput results with high accuracy. However, currently, the technology is cost-intensive and offers limited clinical use in resource-poor settings. In this study, we adopted ELISA, CBA array and qRT-PCR-based validation in independent validation sets, and these methods concurred with the bio-signatures discovered in the Olink experiment. Together, we have introduced a workflow that enables conventional technologies to seamlessly integrate with high-end discovery platforms.

There are a few limitations of this study. The serum protein abundances were monitored at the time of diagnosis and at the completion of therapeutic intervention; however, to capture the dynamics of the inflammatory response in neonatal sepsis, multiple time point sampling will be useful. Validation could be attempted from a larger population, and diverse clinical sites and countries. A second limitation is that our qRT-PCR and immunoplatform-based analysis targeted only a limited number of cytokines. In the future, unbiased discovery-based proteomics and expansion of targets in our downstream analysis would add more rigour to our results. Third, the same patient groups could be used to profile cytokines and other bio-signatures to identify specific correlations in neonatal sepsis. To the best of our knowledge, this is the first report of a multiomics analysis of neonatal venous blood in sepsis, which could help subclassify neonatal sepsis.

## Conclusion

Sepsis is a leading cause of morbidity and mortality in neonates, and identification of the markers contributing to their immune dysregulation might be useful for early disease diagnosis. This study demonstrates an inflammatory biosignature of neonatal sepsis that can differentiate between different sepsis subgroups and from healthy controls. This biosignature of neonatal sepsis is characterized by increased serum pro-inflammatory cytokines such as IL- 6, LIF, and TNF-α levels and decreased IFN-γ and CCL28 levels, which are linked to the serum iron, selenium and copper levels. In addition to the commonly used serum IL-6 as a marker for sepsis diagnosis, other cytokines such as IL-10, also contribute as the central mediator of the inflammatory cascade and a possible recovery marker. This study identified a putative serum biosignature that explains the inflammatory landscape in neonatal sepsis, paving the way for better diagnostic and classification strategies.

## Supporting Information

Supplementary Figure S1: Serum inflammatory proteins show differential expression in neonatal sepsis cases compared with the No Sepsis (NS) group.

Supplementary Figure S2: Serum elemental composition of sepsis patients in different study groups (validation set).

Supplementary Figure S3: Plasma cytokine profiles in neonatal clinical cohorts.

Supplementary Table S1: Epidemiological and clinical details of the neonatal sepsis and control subjects used in this study.

Supplementary Table S2: The list of serum proteins quantified using Olink® Target 92 Inflammation panel.

Supplementary Table S3: Olink proteomics data: NPX protein expression values in wide format.

## Author contributions

Conceptualization: R.N., D.G. Experiment design: R.N., S.T., P.G., D.G. Methodology: D.G., P.G., S.T., R.A. Clinical samples and data: P.D., R.G., G.P.K., R.G., K.N., R.K., S.N., V.K. Sample storage, processing and record keeping: R.A., S.T., A.R, P.G. Clinical data management and analysis: R.A., S.T., A.R. Sample processing: S.T., V.S.N., P.D., P.G, A.R. Data acquisition: S.T., V.S.N., P.D., P.G, A.R, D.G. Data analysis: S.T., V.S.N., P.G., A.R. Investigation: S.T., P.G. Supervision: R.N., M.J.S., D.G. Funding acquisition: R.N., R.G., R.K., S.N., G.P.K., R.G., K.N., D.G., M.J.S. Coordination and strategy: R.N., M.J.S., D.G., R.A. Writing original draft: S.T., D.G, P.G, and R.N. Writing-review and editing: S.T., P.G, V.S.N, R.G., R.K., S.N., G.P.K., R.G., K.N., V.K., D.K., A.C., K.A., R.A., M.J.S., D.G. R.N. All authors read and approved the final manuscript and had full access to all the data in the study.

## Data Availability

All data produced in the present study are available upon reasonable request to the author

## Acknowledgements

We thank the hospital staff at the clinical sites for leading the sample collection and pre-processing. We would like to acknowledge Dr(s) Sushil Shrivastava, Meetu Salhan, Harsh Chellani, Pratima Anand, and Narendra Pal Singh for their constant support and involvement in this project. We would also like to thank the Department of Biotechnology (DBT), India, for financial support. We gratefully acknowledge Olink^®^ Proteomics AB, Sweden, for providing technical support. We also thank the Translational Health group members for their help and support.

