## Supplementary File for "Serum IL-6, IL-10, TNF-α and IFN-γ bio-signature for neonatal sepsis diagnosis and treatment outcome"

| **TABLE OF CONTENTS** | |
| --- | --- |
| **Supplementary Figure S1:** | Serum inflammatory proteins show differential expression in neonatal sepsis cases compared with the No Sepsis (NS) group. |
| **Supplementary Figure S2:** | Serum elemental composition of sepsis patients in different study groups (validation set). |
| **Supplementary Figure S3:** | Plasma cytokine profiles in neonatal clinical cohorts |
| **Supplementary Table S1:** | The Table containing the clinical details of the neonatal sepsis and control study subjects used in this study is presented in an Excel file (Supplementary Table S1). |
| **Supplementary Table S2:** | The list of 92 serum proteins quantified using the Olink® Target 92 inflammation panel. |
| **Supplementary Table S3:** | Olink proteomics data: NPX protein expression values in wide format are presented in a separate Excel file (Supplementary Table S3). |

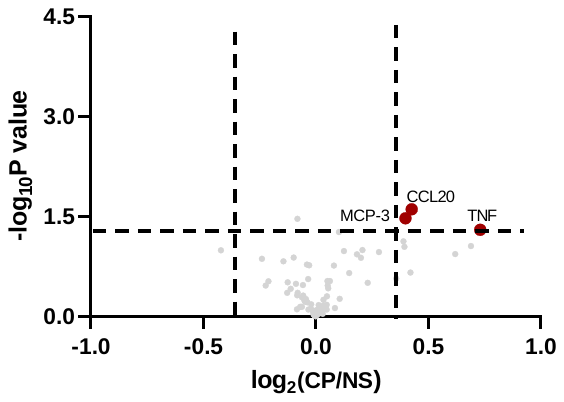

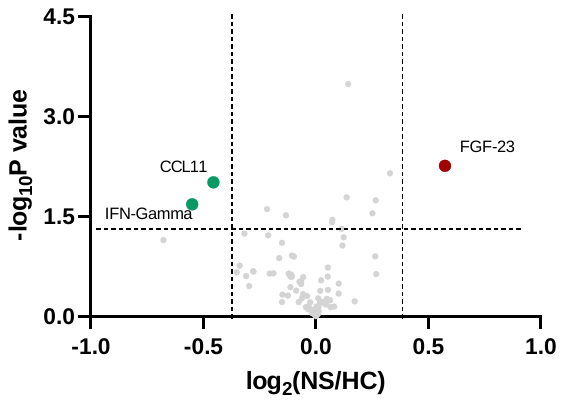

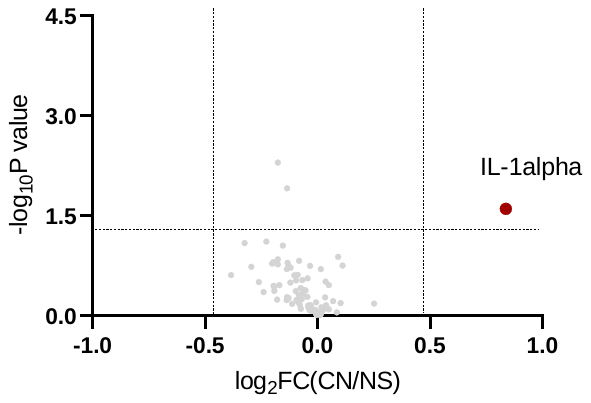

A

B

C

**Supplementary Figure S1. Serum inflammatory proteins show differential expression in neonatal sepsis cases compared with the No Sepsis (NS) group.** Volcano plots showing dysregulated proteins (log_2_(FC)>±1.3, p-value < 0.05) between study groups, **A.** culture positive (CP), **B.** culture negative (CN) neonatal sepsis groups. **C.** Healthy (HC) and No sepsis (NS) controls.

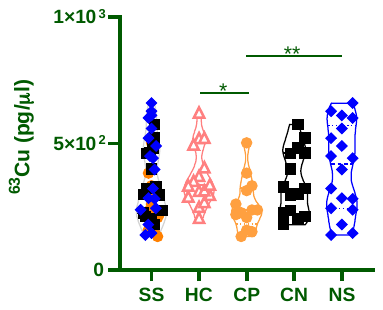

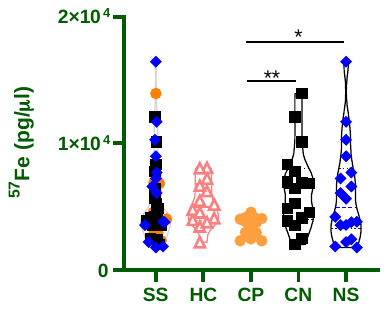

A

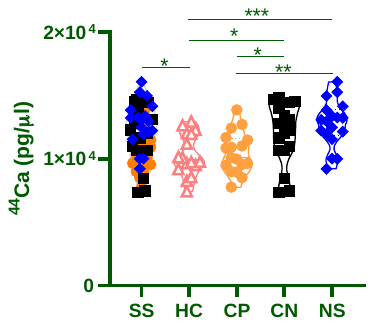

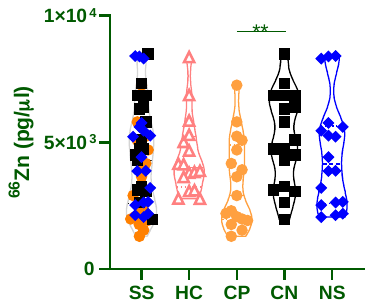

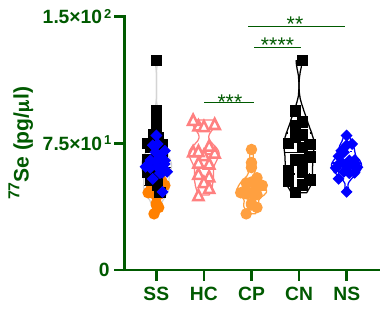

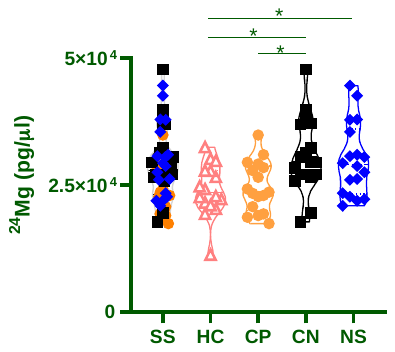

**Supplementary Figure S2. Serum elemental composition of sepsis patients in different study groups (validation set). A.** Violin plots of serum Iron, Copper, Calcium, Magnesium, Zinc, and Selenium between culture positive (CP), culture negative (CN), no-sepsis (NS) and healthy controls (HC). (p-value <0.05:*,<0.005:**, <0.0005: ***, <0.0001:****).

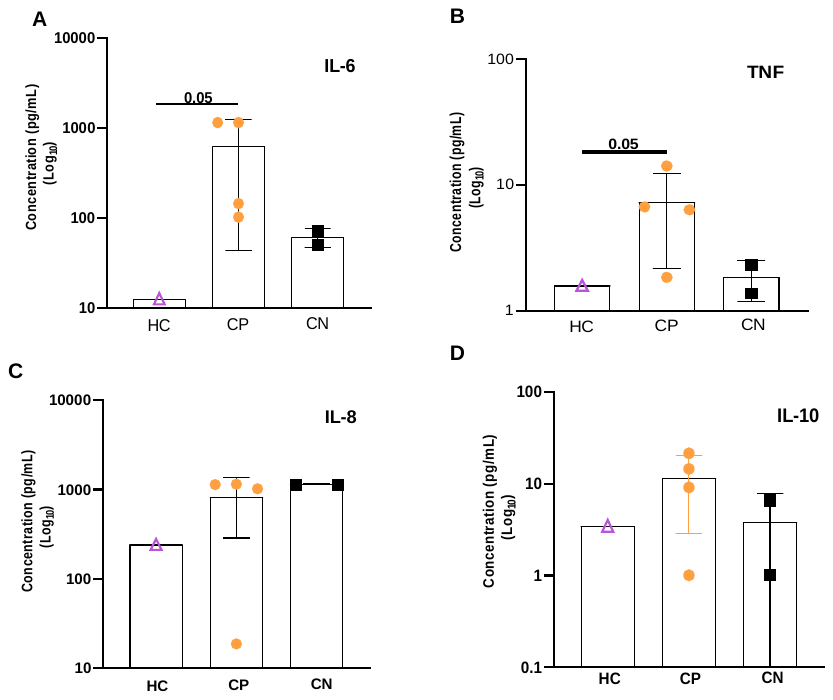

**Supplementary Figure S3. Plasma cytokine profiles in neonatal clinical cohorts.** Plasma concentrations of (A) TNF, (B) IL-6, (C) IL-8, and (D) IL-10 were quantified using a Cytometric Bead Array (CBA) across No sepsis (NS), Culture-positive (CP), and Culture-negative (CN) sepsis cohorts. Cytokine concentrations are expressed in pg/mL and plotted on a Log10 scale. Individual data points represent the mean of technical replicates for each neonate, while bars indicate the cohort mean ± SEM. Statistical significance was evaluated using a One-way Analysis of Variance (ANOVA) followed by Dunnett’s multiple comparisons test against the NS baseline cohort. Significance was defined as p < 0.05.

**Supplementary Table S1. The Table containing the clinical details of the neonatal sepsis and control study subjects used in this study is presented in an Excel file (Supplementary Table S1).**

**Supplementary Table S2. The list of 92 serum proteins quantified using the Olink® Target 92 inflammation panel.**

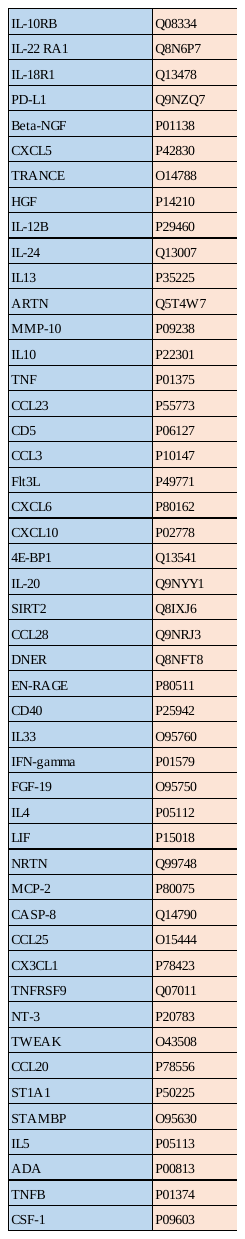

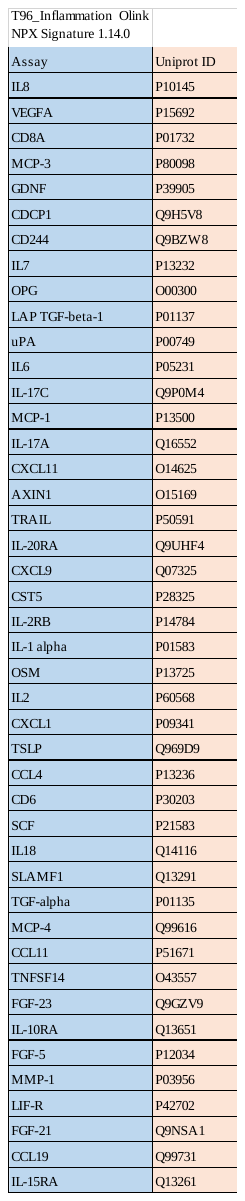

**Supplementary Table S3: Olink proteomics data: NPX protein expression values in wide format are presented in a separate Excel file (Supplementary Table S3).**
